# How to quantify deaths averted derived from interrupted time-series analyses

**DOI:** 10.1101/2021.03.23.21254181

**Authors:** Huan Jiang, Alexander Tran, Gerhard Gmel, Shannon Lange, Jakob Manthey, Robin Room, Pol Rovira, Mindaugas Štelemėkas, Tadas Telksnys, Jürgen Rehm

## Abstract

**Background:** Interrupted time series (ITS) are an important tool for determining whether alcohol control policies, as well as other policy interventions, are successful over and above secular trends or chance. Subsequent to estimating whether a policy has had an effect, quantifying the key outcomes, such as the number of prevented deaths, is of primary practical importance. The current paper compares the results of two different methodological approaches to quantify deaths averted using different two standard populations.

**Methods:** Time series methodologies were used to estimate the effect size in deaths averted of a substantial increase in excise taxation in Lithuania in 2017. We compare the impact of a) using ITS methodology vs. fitting the trend before the intervention to predict the following 12 months and comparing the predicted monthly estimates of deaths with the actual numbers; and b) adjusting the time series either using the World Health Organization standard or the age distribution of Lithuania in the month before the intervention. The effect was estimated by sex.

**Results:** The increase in excise taxation was associated with a substantial decrease in all-cause mortality in all models considered. ITS methodology and using the age-distribution of Lithuania were consistently associated with higher estimates of deaths averted. Although confidence and prediction intervals were highly overlapping, the point estimates differed substantially. The taxation increase was associated with 1,155 deaths averted in the year following the intervention (95% prediction interval: 729, 1,582), corresponding to 2.80% of all deaths in Lithuania in the respective year, for the model selected as best for planning policy interventions in Lithuania.

**Conclusions:** Fitting a time series model for the time until the intervention, and then comparing the predicted time points with the actual mortality, standardizing to country-specific weights, was chosen as the best way to derive practically relevant effect sizes.

## Introduction

Alcohol use has been identified as a major risk factor for the global burden of mortality and morbidity in all comparative risk analyses to date ((Rehm and Imtiaz, 2016); for latest efforts, see (Shield et al., 2020, GBD 2019 Risk Factors Collaborators, 2020, World Health Organization, 2018)). Fortunately, effective evidence-based alcohol control policies, such as the World Health Organization’s (WHO) “best buys” for alcohol (increase in taxation, reduction of availability of alcoholic beverages, and a ban on advertisement) (World Health Organization, 2017), can protect population health and save lives, thereby reducing health care costs and increasing productivity (Babor, 2010).

Given that the impact of regulatory and public policy initiatives cannot usually be tested through traditional randomized controlled trial designs, well-selected, -designed, and -analyzed natural experiments are the method of choice when examining the effects of such enactments on a variety of outcomes (Craig et al., 2017, Dunning, 2012, Shadish et al., 2002). The classic methodology for such evaluations is interrupted time-series analysis (ITS), which is considered as one of the quasi-experimental designs that use both pre- and post-policy data without randomization (Beard et al., 2019). Subsequent to estimating whether a policy has had an effect over and above secular trends and chance (Shadish et al., 2002), a quantification of the effect size in tangible units, such as the number of prevented cases/deaths, often is of primary practical importance, as this information is key for future decisions aiming to maximize public health promotion and mitigate alcohol-related harms. In time-series analyses, there are two main types of effect-size estimates that describe the impact of the policy: first, level change, and second, slope change (Bernal et al., 2016).

In public health, the estimation of the number of prevented cases/deaths should take age into consideration, as most health indicators such as mortality rate or prevalence of disease are highly associated with age. However, the population structure often changes during the period used in the time-series analysis to evaluate the impact of an intervention, so age-standardization, i.e., weighting age-specific rates based on a synthetic or other standard population (Statistics Canada, 2020) is used to make the time points comparable. However, an estimate of deaths based on a standard population weighted heavily towards older age groups would result in a higher number of deaths, while an estimate of deaths based on a standard population weighted towards younger people would shift the results toward a lower number of deaths. Thus, the estimated effect of an intervention policy may be impacted by the standard population chosen.

In the current paper, we will focus on the translation of results from ITS into tangible results such as deaths averted. We will compare the impact of two different methodological approaches to quantify deaths averted using different standard populations, resulting in four different estimates. The example used will be the implementation of a substantial increase in excise taxes in Lithuania in 2017 and its effect on the reduction of adult (20+ years of age) all-cause mortality. In detail, on March 1, 2017, there were marked increases in excise taxes - 111-112% for fermented beverages, wines and beer; 91-94% for intermediate products (such as fortified wines); and 23% for ethyl alcohol (applying for spirits) - reducing the affordability of alcohol (Rehm et al., 2021). As shown elsewhere, this alcohol control policy resulted in a substantial decrease in alcohol mortality rates, especially for men (Štelemėkas et al., 2021). The aim of this paper will be to quantify this effect into deaths averted in the year after its implementation, based on different methodologies.

## Methods

### Time of investigation and data sources

We used month-by-month age-standardized sex-specific mortality rates for Lithuania from January 2000 to December 2019 as the basis for the ITS. The number of deaths was obtained from the Lithuanian Institute of Hygiene, and the population breakdown by sex and age was obtained from Statistics Lithuania (Lithuanian Department of Statistics, 2021).

### Age-standardization

In order to age-standardize all-cause mortality rates for people over 20 years of age, the age-specific Lithuanian mortality rates (in 10-year-intervals) were weighted by 1) the WHO global standard (Ahmad et al., 2001), and 2) the age distribution of the Lithuanian population in February 2017, the month before the taxation increase (obtained from Statistics Lithuania (Lithuanian Department of Statistics, 2021).

### Estimating the Intervention Effect

The time-series analyses were conducted using generalized additive models (GAMs (Beard et al., 2019, Hastie and Tibshirani, 1990); see also description in the Appendix), which were adjusted for seasonality by smoothing splines. The association of all-cause mortality with the increase in excise taxes for alcohol was our main interest, but for comparison reasons we based our analyses on the paper of Štelemėkas and colleagues (Štelemėkas et al., 2021), where several policies were compared, and thus entered the same set into the model (for selection of policies, see (Rehm et al., 2021)). All policies were coded as abrupt permanent effects, i.e., assigning a value of 0 for all months preceding the implementation and a value of 1 for months after. Potential confounding factors included centered monthly gross domestic product (GDP), centered unemployment rate, and a second-order polynomial of the time to account for the overall curvilinear trend in all-cause mortality. The final models were determined based on the following criteria: a) adjustment for at least one economic indicator (GDP or unemployment rate); b) independence and presence of non-stationarity in residuals (checked with residual plots and confirmed by the augmented Dickey-Fuller test); c) model-fitting statistics (AIC and BIC) and likelihood ratio test of nested models for superior models.

The policy intervention effect was estimated with two different methods. (1) We applied the standard way of a simple multiplication of the effect of the intervention in the ITS, where the beta-weight for the effect on the age-standardized mortality rate is multiplied by the average population for the 12 months following the intervention. (2) For the second method, we followed a three-step approach:

**Step 1:** Use the data **before** the intervention point to determine the optimal GAM model for the series.

**Step 2:** Use that GAM model to forecast mortality rates for 12 months after the intervention. For each month, the forecasted mortality rates and their 95% prediction intervals (PIs) were calculated assuming that the forecast errors were normally distributed.

**Step 3:** The differences in mortality rates between actual values and forecast values after the intervention were multiplied by the corresponding population, resulting in an estimate of the absolute numbers of deaths being averted for each month. The sum of those absolute numbers was considered to be the estimated averted death for the year after the intervention, whose standard deviation was calculated by taking the square root of the combined variances, assuming independence between monthly data points. The 95% PI was further calculated assuming the forecast errors being normally distributed.

Both methods were applied to women and men separately. The effect for the total population was estimated in two ways. First, by adding the results for women and men together (as reported in the Tables), and, second, for a sensitivity analysis by estimating the effect based on the time series of the total mortality rate. We used two underlying standard populations: the WHO standard population, which is based on the global age distribution with higher weights for younger age groups ((Ahmad et al., 2001); 87.41% of the age group 20 and older between ages 20 and 64), and the Lithuanian population distribution of the month before the intervention ((Lithuanian Department of Statistics, 2021); 75.79% of the age group 20 and older are between ages 20 and 64)). Both age-distributions used as a standard can be found in the Appendix Table A.4.

All analyses were performed using R version 4.0.4. (The Comprehensive R Archive Network (CRAN), 2021).

## Results

Overall, all-cause mortality decreased during the period 2001 to 2018 (see Appendix Table A.1). As reported in detail elsewhere (Štelemėkas et al., 2021), when using the WHO standard population, the effect size of the 2017 policy was estimated to be −11.07 (95% CI: − 18.88, −3.37) for men, compared to an estimate of −14.16 (95% CI: −26.65, −1.67) using the Lithuania population as of February 2017 as the standard (see Appendix Table A.4 for details). Simply applying these effect sizes to the average population of Lithuania in the year after the taxation increase (March 2017 to February 2018) resulted in 1,344 (95% CI: 397, 2,290), and 1,718 (95% CI: 203, 3,232) averted deaths among men, respectively (Table 1).

**Table 1.** ITS analysis: estimated effect sizes and confidence intervals for the period March 2017 – February 2018. The results for the total population were based on the sex-specific results.

| Standard Population |  | Women |  | Men |  | Total |  |
| --- | --- | --- | --- | --- | --- | --- | --- |
|  |  | Effect size | Averted deaths | Effect size | Averted deaths | Effect size | Averted deaths |
| WHO standard population | Estimate | -2.29 | 343 | -11.07 | 1,344 | -6.23 | 1,687 |
|  | 95% CI | (-6.17, 1.58) | (-235, 920) | (-18.88, -3.27) | (397, 2,290) | (-10.33, -2.12) | (578, 2,795) |
| Lithuanian population 2017 | Estimate | -2.90 | 433 | -14.16 | 1,718 | -7.95 | 2,151 |
|  | 95% CI | (-9.56, 3.75) | (-560, 1427) | (-26.65, -1.67) | (203, 3,232) | (-14.65, -1.25) | (340, 3,962) |

When using the above-described methodology of fitting a time-series analysis until February 2017, and then deriving the number of deaths averted by subtracting the estimated deaths in each of the 12 months post-intervention (i.e., until and including February 2018) from the real number of deaths, the following results could be derived: 1) based on the WHO standard population, the averted number of deaths among men was estimated to be 864(95% PI: 651, 1077), while 2) using the Lithuania population of 2017 as the standard, the averted number of deaths among men was estimated to be 979(95% PI: 618, 1340). Figure 1 and 2 show the predicted monthly all-cause mortality rates and the observed rates for men for the 12 months following the taxation increases using the two different age standardization schemes (the respective figures for women can be found in the Appendix).

**Figure 1.**
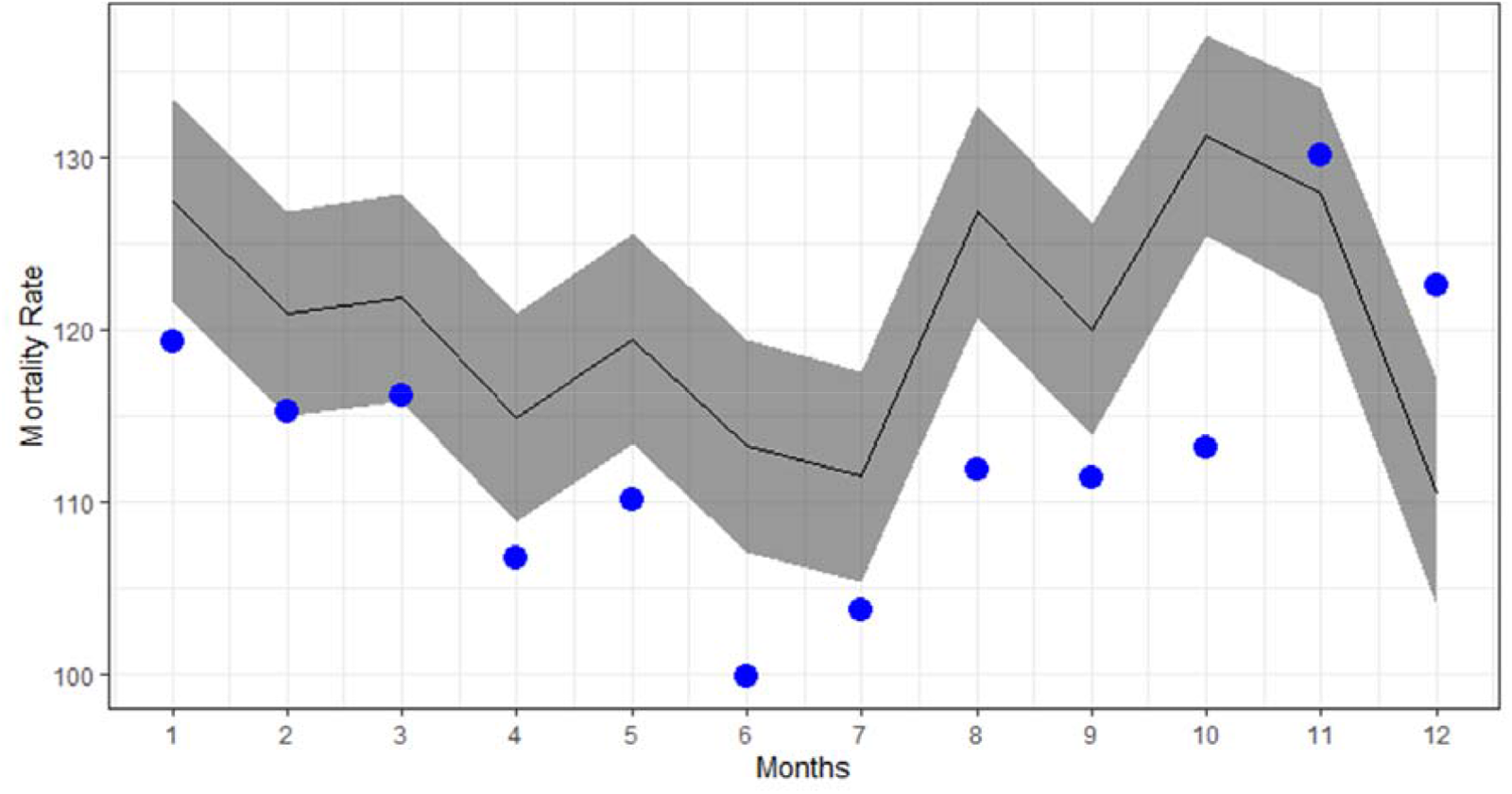
Using the World Health Organization standard population: predicted and observed all-cause mortality rates for men for the period March 2017 – February 2018 in Lithuania. Black lines indicate the forecasted number of deaths without the 2017 taxation intervention. Blue dots indicate observed deaths.

**Figure 2.**
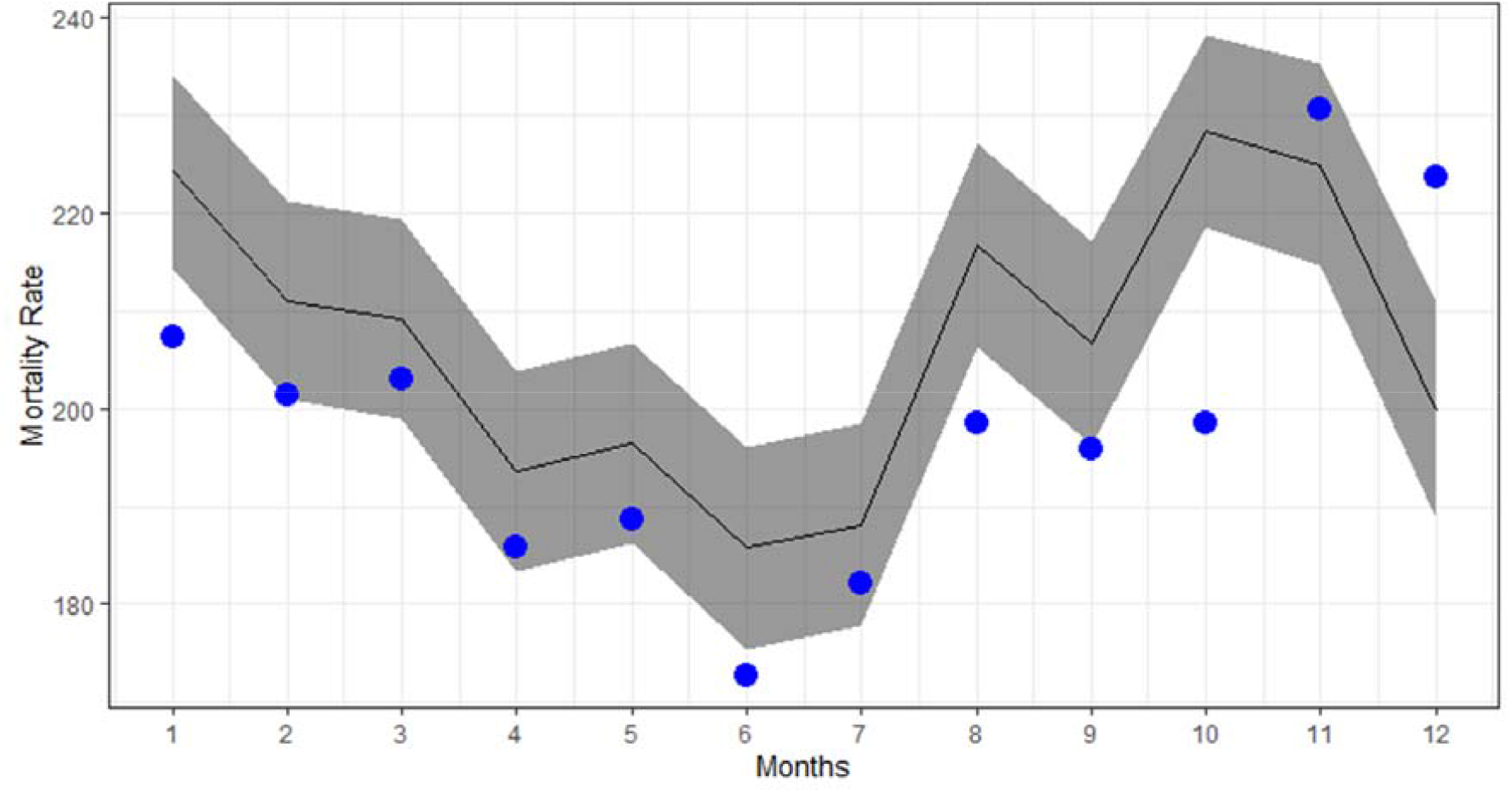
Using Lithuanian population as the standard: predicted and observed all-cause mortality rates for men for the period March 2017 – February 2018 in Lithuania. Black lines indicate the forecasted number of deaths without the 2017 taxation intervention. Blue dots indicate observed deaths.

The results for women and both sexes combined based on the ITS analyses with estimates of averted mortality being derived from the beta weights are summarized in Table 1 and Table 2, along with the results for men. Details of the underlying models can be found in the Appendix Table A.2, A.3, and in the publication of Štelemėkas and colleagues, already mentioned above (Štelemėkas et al., 2021).

**Table 2.** Prediction analyses: estimates of averted number of deaths and their prediction interval (PI) for the period of March 2017 – February 2018, based on a three-step approach. The results for the total population were based on the sex-specific results.

| Standard Population |  | Women | Men | Total |
| --- | --- | --- | --- | --- |
| WHO standard population | Estimate | 172 | 864 | 1,036 |
|  | 95% PI | (58, 286) | (651, 1,077) | (794, 1,278) |
| Lithuanian population 2017 | Estimate | 176 | 979 | 1,155 |
|  | 95% PI | (-51, 404) | (618, 1,340) | (729, 1,582) |

The numbers of averted deaths for men, 1344 and 1718 deaths within a 1-year period, depending on the standard population used, correspond to 6.64% and 8.48% of the total male mortality in the prior year. For women, the proportions of death averted amounted to 1.64% and 2.07% based on the WHO standard population and the Lithuanian population standard, respectively.

The proportional reductions from the prior year were 4.27% and 4.83% for men, and 0.82% and 0.84% for women for the WHO and Lithuanian standard population, respectively. The sensitivity analysis of a separate ITS of the total population rate corroborated the data: WHO standard averted deaths 949; 95% CI: 612; 1285; Lithuanian standard: 1202; 95% CI: 577-1811.

In sum, we found sizable decreases in all-cause mortality following the intervention with all methodologies, but the point estimates vary by almost a factor of two.

## Discussion

Overall, the results of the models presented here support the findings of other studies on the effects of alcohol control policies ((Neufeld et al., 2021); for the impact of taxation in general, see (Sornpaisarn et al., 2017)), in that all analyses conducted suggest a sizable effect of taxation increases on all-cause mortality. The different methodologies resulted in point estimates that were substantially different, albeit with highly overlapping confidence intervals. Standardization to the Lithuanian population resulted in higher estimates than the standardization to the WHO population, and the more complex comparison of the predicted mortality rate with the real numbers resulted in lower estimates than just applying the actual population to the beta-weights (see below for a discussion).

While the effect of the taxation increase was sizable under all scenarios, it needs to be reiterated that any effects of taxation legislation are, of course, dependent on the size of the tax increase; for Lithuania, the tax increase led to a clear decrease in affordability, which is the prerequisite for a sizable impact on alcohol consumption and harm (Neufeld et al., 2021, Grigoriev and Bobrova, 2020, Herttua et al., 2017), given the underlying economic theory of supply and demand (Sornpaisarn et al., 2017). Too often alcohol tax increases do not result in decreased affordability, and rarely consider inflation and increases in purchasing power. As a consequence, alcoholic beverages have become more affordable in many countries—not just European countries—over the past decades (Blecher et al., 2018, Rabinovich et al., 2009). However, when applied correctly, alcohol taxation constitutes a powerful tool that can result in sizable all-cause mortality reductions.

There are some potential limitations of the current study that should be acknowledged. First, as with all ITS analyses, where we conclude a causal effect of a policy or another event, we need to assume that no other important policy change or relevant event happened on March 1 2017, or close to this date, which could potentially explain the effects (Shadish et al., 2002). None of the Lithuanian experts consulted could list any such event, but we cannot exclude this possibility in an ecological design. In terms of policy changes, even though there were no additional alcohol policies implemented up until the end of 2017, several new alcohol control policies (an alcohol advertising ban; an increase in the legal alcohol purchase age; and additional restriction on sale hours) came into effect on January 1, 2018, and therefore overlap in effects in two of the twelve months following the evaluated tax changes may have impacted the estimates. However, these policy effects were included as independent variables in the ITS and did not contribute to a further decrease in all-cause mortality (see Figure 1 and 2; see also Appendix Table A2). Second, our results assume that the time series modelling until February 2017 represents a good approximation of the actual mortality rates. In our example, this appears to be the case, as our models explained around 80% of the variation (see (Štelemėkas et al., 2021) and Appendix). However, there is inherently some error in our estimates given that these statistical models are based on various assumptions. The major difficulty is that the PIs might be too narrow as a result of not accounting for all sources of uncertainty. For example, we assumed the predicted monthly mortality numbers were independent when constructing the PI, which may well not be accurate. It would thus be prudent to try different models with different assumptions, e.g., Bayesian models (de Vocht et al., 2020, de Vocht et al., 2017), and compare the results. Finally, this work focused on quantifying the deaths averted as a result by one policy intervention. In order to compare to published literature, we replicated a recent model from the literature which tried to measure the effects of several interventions. This may also have introduced bias.

Regardless, from the analyses presented, it may be concluded that comparing the actual number of deaths in each month to the forecasted number gives a more detailed and concrete estimate of the difference attributable to the intervention than using the beta-weight of the ITS analysis, which only gives a rough estimate. The higher estimated number of deaths averted by the classic ITS model may be the result of the modelling strategy, which assumed the same effect size for the remainder of time period, i.e., for longer than one year. In other words: the ITS and the beta-weight method uses one model to describe the whole time series and assumes the continuation of the historical data-generating process over this period. On the contrary, the three-step approach uses a model to describe the historical time series before the intervention and potentially allows for different processes generating the data after the intervention. Therefore, it should be the preferred approach compared to the beta-weight method, if the objective is to quantify the size of the effect.

As for age-standardization, clearly the standard used made a sizable difference. Deciding on which standard should be used would depend on the objective of the research. If we want to compare across countries or globally, it is best to use the WHO standard (Ahmad et al., 2001), which is used in most comparative publications including the Global Burden of Disease Study (GBD 2019 Risk Factors Collaborators, 2020). However, if the objective is to get the best estimate for a specific country, in this case the number of deaths averted in Lithuania after the policy intervention of March 1, 2017, it is best to age-standardize based on a recent age-distribution for that country. With that being said, it should always be kept in mind that age-standardized rates are a hypothetical construct and should not be interpreted as reflecting actual risk. The use of a recent country-specific standard will lessen the extent of any such misinterpretation when evaluating policies in years close to those used in the standard.

Finally, we return briefly to the implications of the Lithuanian experience, and of the particular intervention studied in this paper, for the World Health Organization’s global public health strategy to reduce harms from alcohol, and its identification of “best buys” for national alcohol policies (World Health Organization, 2017). Based on our best estimates using the Lithuanian age distribution, reducing the overall adult mortality in a high-income country like Lithuania by 4.83% for men, 0.84% for women and 2.80% overall is a great achievement for a single intervention, and truly deserves the label of a “best buy”.

## Data Availability

The data that support the findings of this study are available from the corresponding author, HJ, upon reasonable request.

## Acknowledgements

JR acknowledges funding from the Canadian Institutes of Health Research, Institute of Neurosciences, and Mental Health and Addiction (CRISM Ontario Node grant no. SMN-13950). Research reported in this publication was also supported by the National Institute on Alcohol Abuse and Alcoholism of the National Institutes of Health (NIAAA) [Award Number 1R01AA028224-01].

Content is the responsibility of the authors and does not reflect official positions of NIAAA or the National Institutes of Health.

The authors have no conflict of interest to declare.

## Acknowledgements

We would like to thank Ms. Astrid Otto for referencing and copy-editing the text.

## Appendices

### Generalized Additive Models (GAMs)

GAM model structure (Hastie and Tibshirani, 1990): *g*(*E*(*Y*))=*α*+*s*_1_(*x*_1_)+…+*s*_*p*_(*x*_*p*_) where ***Y*** is the dependent variable, ***g*(*Y*)** denotes the link function, and the terms *s*_*i*_(*x*_*i*_) denote smooth, non-parametric functions. The terminology “nonparametric” means that the shape of predictor functions are fully driven by the data as opposed to parametric functions that are defined by a typically small set of parameters from a certain distribution. This allow for more flexible estimation of the underlying predictive patterns without knowing upfront what these patterns look like. When there is substantial residual autocorrelation in the data, an ARMA process is fit to the residuals.

**Table A.1.**
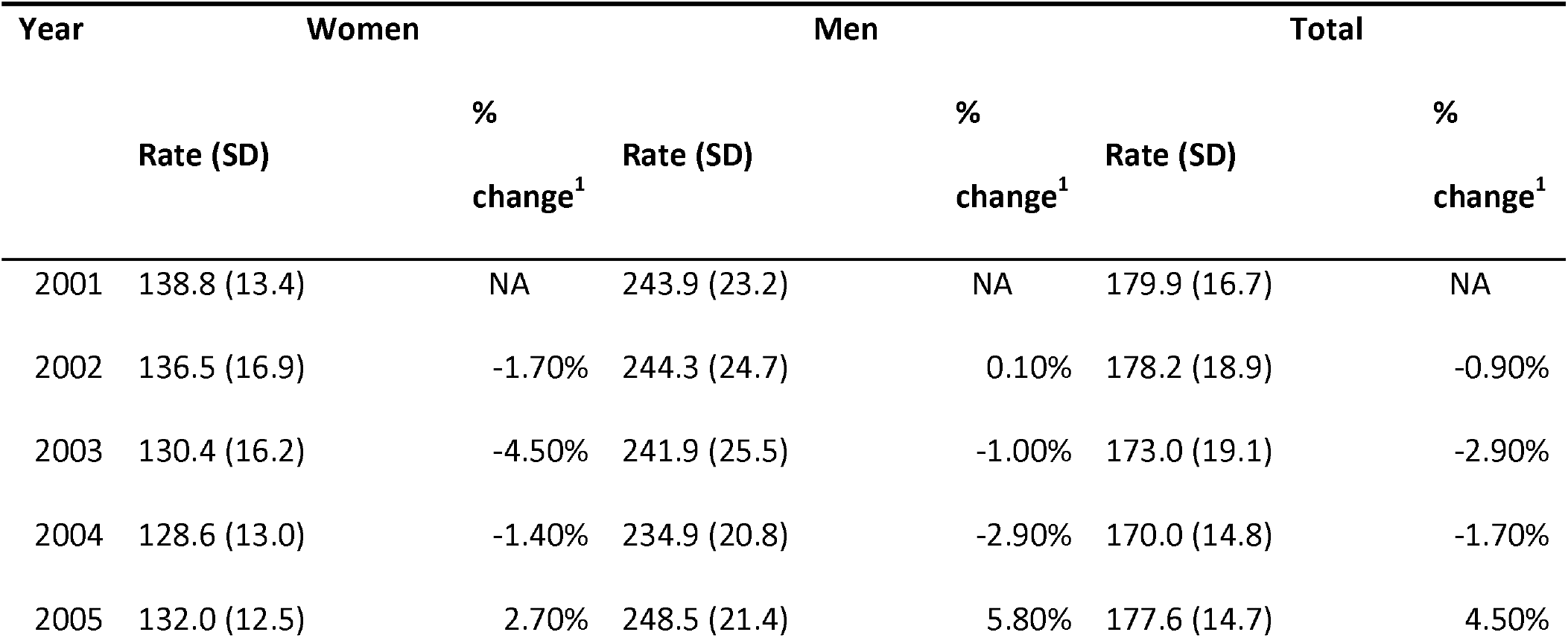
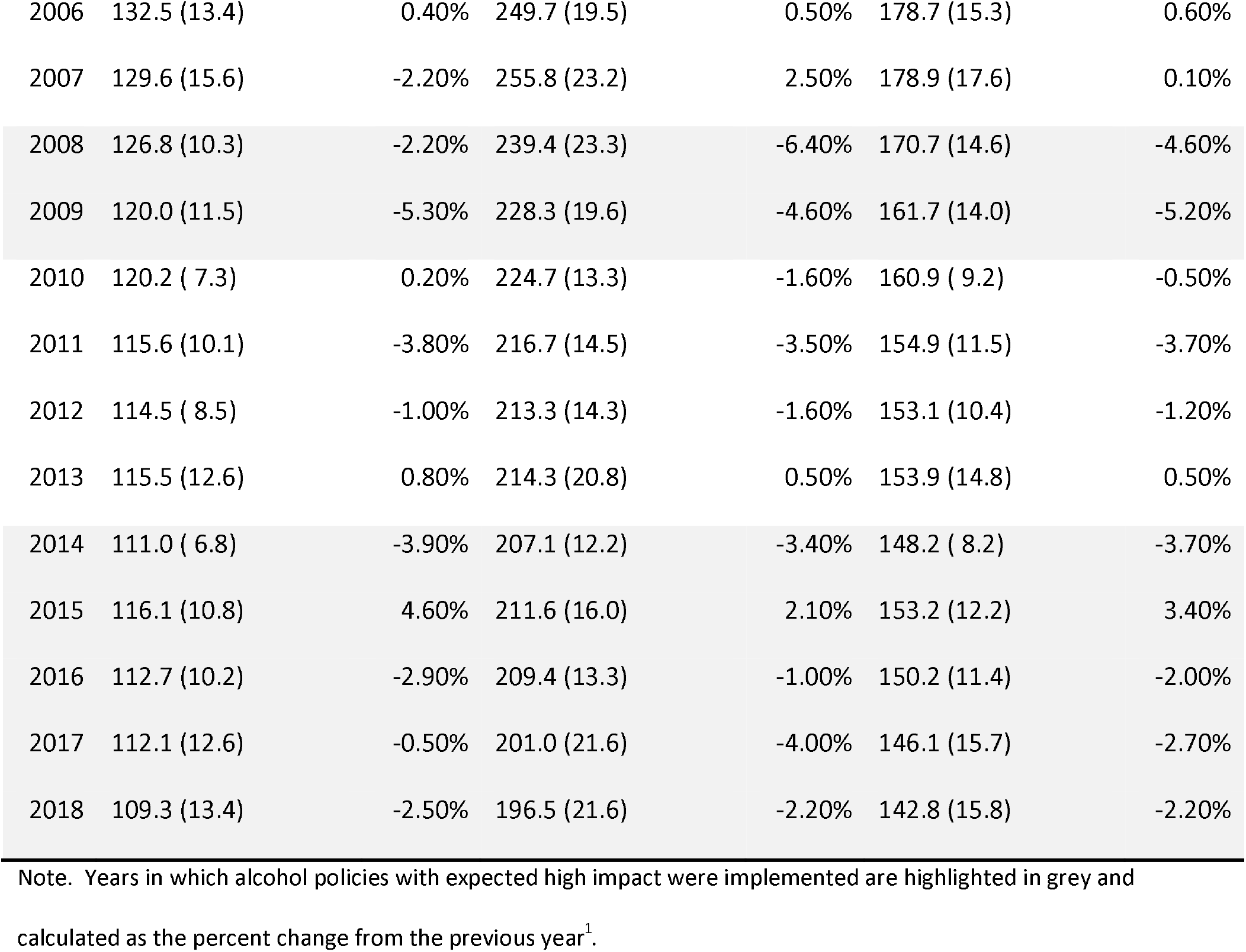
Sex-stratified and total annual age-standardized all-cause mortality rates per 100,000, using the Lithuanian population from February 2017 as the standard population

**Table A.2.** Model results for the impact of different sets of alcohol policies on all-cause mortality, using the Lithuanian population from February 2017 as the standard population

|  | Women | Men | Total |
| --- | --- | --- | --- |
| Intercept | 126.30 (122.76, 129.83)*** | 271.81 (257.74, 285.88)*** | 185.57 (175.37, 195.78)*** |
| Months^2 |  | -0.003 (-0.004, -0.002)*** | -0.001 (-0.002, -0.0004)*** |
| GDP *per capita* in 1,000s<br>Euros (centered) | -7.19 (-10.58, -3.80)*** | 21.13 (9.05, 33.21)*** | 5.84 (-2.93, 14.62) |
| Policy implementation date<sup>4</sup> |  |  |  |
| 1-Jan-08 | 1.70 (-4.41, 7.81) | -10.91 (-22.95, 1.14)* | -3.80 (-12.62, 5.01) |
| 1-Jan-09 | -10.23 (-15.27, -5.19)*** | 1.37 (-13.17, 15.91) | -3.76 (-14.34, 6.83) |
| 1-Apr-14 | 1.59 (-3.85, 7.04) | 7.85 (-4.86, 20.57) | 5.87 (-3.26, 15.00) |
| 1-Mar-15 | 2.05 (-4.81, 8.91) | 11.03 (-2.03, 24.10)* | 6.00 (-3.39, 15.40) |
| 1-Jan-16 | 0.71 (-5.92, 7.33) | 10.75 (-2.49, 23.99) | 6.35 (-3.32, 16.02) |
| 1-Mar-17 | -2.90 (-9.56, 3.75) | -14.16 (-26.65, -1.67)** | -8.62 (-17.63, 0.39)* |
| 1-Jan-18 | 2.09 (-4.82, 9.00) | 9.92 (-3.59, 23.42) | 6.84 (-3.04, 16.71) |
| R<sup>2</sup> | 0.80 | 0.82 | 0.83 |
| Autocorrelation<sup>1</sup> | MA(1) | AR(1) | AR(1) |

Note. \* p<.05 ; \*\* p<.01 ; \*\*\* p<.001.
Results from generalized additive models using the age-standardized all-cause mortality rates per 100,000 people as the dependent variable and adjusted for autocorrelation and seasonality.
<sup>1</sup>The terms used to correct for autocorrelation in the time-series: AR[number] = autoregressive term of order [number], MA[number] = moving average term of order [number]

**Table A.3.** Predictive model results on all-cause mortality for the impact of different sets of alcohol policies before February 2017, using the Lithuanian population from February 2017 as the standard population

|  | Women | Men | Total |
| --- | --- | --- | --- |
| Intercept | 126.47 (122.94, 130.00)*** | 272.45 (257.95, 286.95)*** | 185.95 (174.69, 197.21)*** |
| Months<sup>2</sup> |  | -0.003 (-0.004, -0.001)*** | -0.001 (-0.002, -0.0004)*** |
| GDP *per capita* in 1,000s Euros (centered) | -7.06 (-10.45, -3.67)*** | 21.80 (9.33, 34.26)*** | 6.08 (-3.66, 15.82) |
| Policy implementation date |  |  |  |
| 1-Jan-08 | 1.57 (-4.50, 7.64) | -11.67 (-24.14, 0.81)* | -3.89 (-13.92, 6.14) |
| 1-Jan-09 | -10.20 (-15.19, -5.21)*** | 0.93 (-14.05, 15.91) | -3.82 (-15.64, 8.00) |
| 1-Apr-14 | 1.51 (-3.90, 6.92) | 7.12 (-5.86, 20.09) | 5.81 (-3.93, 15.56) |
| 1-Mar-15 | 2.03 (-4.77, 8.83) | 10.76 (-2.54, 24.06) | 5.71 (-4.33, 15.74) |
| 1-Jan-16 | 0.53 (-6.04, 7.09) | 10.69 (-2.99, 24.36) | 7.17 (-3.74, 18.09) |
| R<sup>2</sup> | 0.78 | 0.79 | 0.81 |
| Autocorrelation<sup>1</sup> | MA(1) | AR(2) | AR(2) |

Note. \* p<.05 ; \*\* p<.01 ; \*\*\* p<.001.
Results from generalized additive models using the age-standardized all-cause mortality rates per 100,000 people as the dependent variable and adjusted for autocorrelation and seasonality.
<sup>1</sup>The terms used to correct for autocorrelation in the time-series: AR[number] = autoregressive term of order [number], MA[number] = moving average term of order [number]

**Table A.4.** Standardization values for World Health Organization Standard and the Lithuanian population in February 2017 (ten-year intervals)

| WHO Standard |  |  | LTU Population (February 2017) |  |  |
| --- | --- | --- | --- | --- | --- |
| Age group | Total Weighting | 20+ Weighting | Age group | Total Weighting | 20+ Weighting |
| 0-9 | 0.17515 | - | 0-9 | 0.102977 | - |
| 10-19 | 0.1707 | - | 10-19 | 0.098923 | - |
| 20-29 | 0.1615 | 0.246885 | 20-29 | 0.131529 | 0.164803 |
| 30-39 | 0.1476 | 0.225636 | 30-39 | 0.122113 | 0.153005 |
| 40-49 | 0.1263 | 0.193075 | 40-49 | 0.136919 | 0.171556 |
| 50-59 | 0.0992 | 0.151647 | 50-59 | 0.153043 | 0.191759 |
| 60-69 | 0.0668 | 0.102117 | 60-69 | 0.11431 | 0.143228 |
| 70-79 | 0.0373 | 0.057021 | 70-79 | 0.085306 | 0.106886 |
| 80+ | 0.01545 | 0.023618 | 80+ | 0.05488 | 0.068763 |

**Figure A.2.**
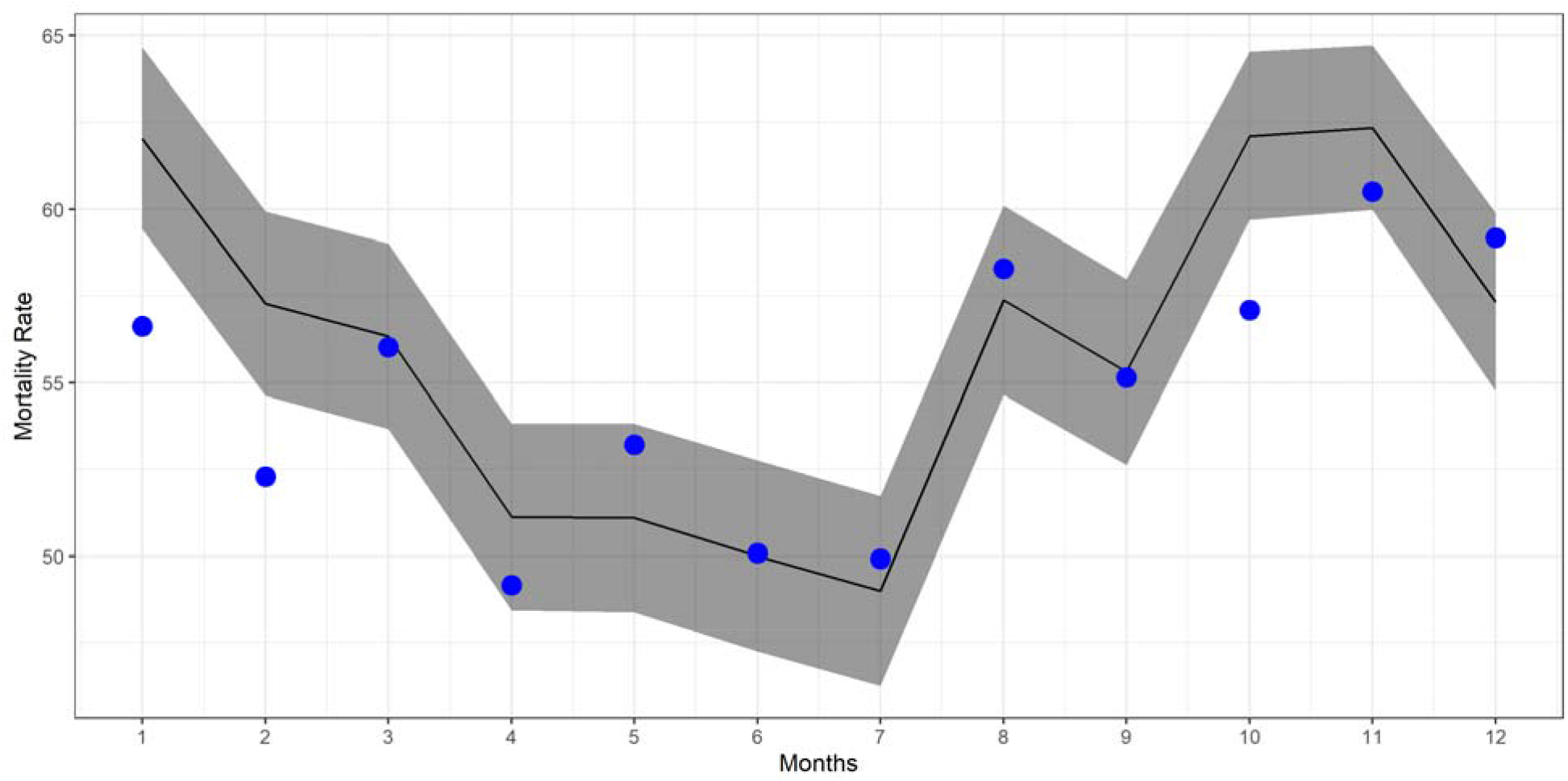
Predicted and observed all-cause mortality rates for females for the period March 2017 – February 2018 in Lithuania. Black lines indicate the forecast number of deaths without the 2017 taxation intervention, using the WHO standard population. Blue dots indicate observed mortality rates.

**Figure A.2.**
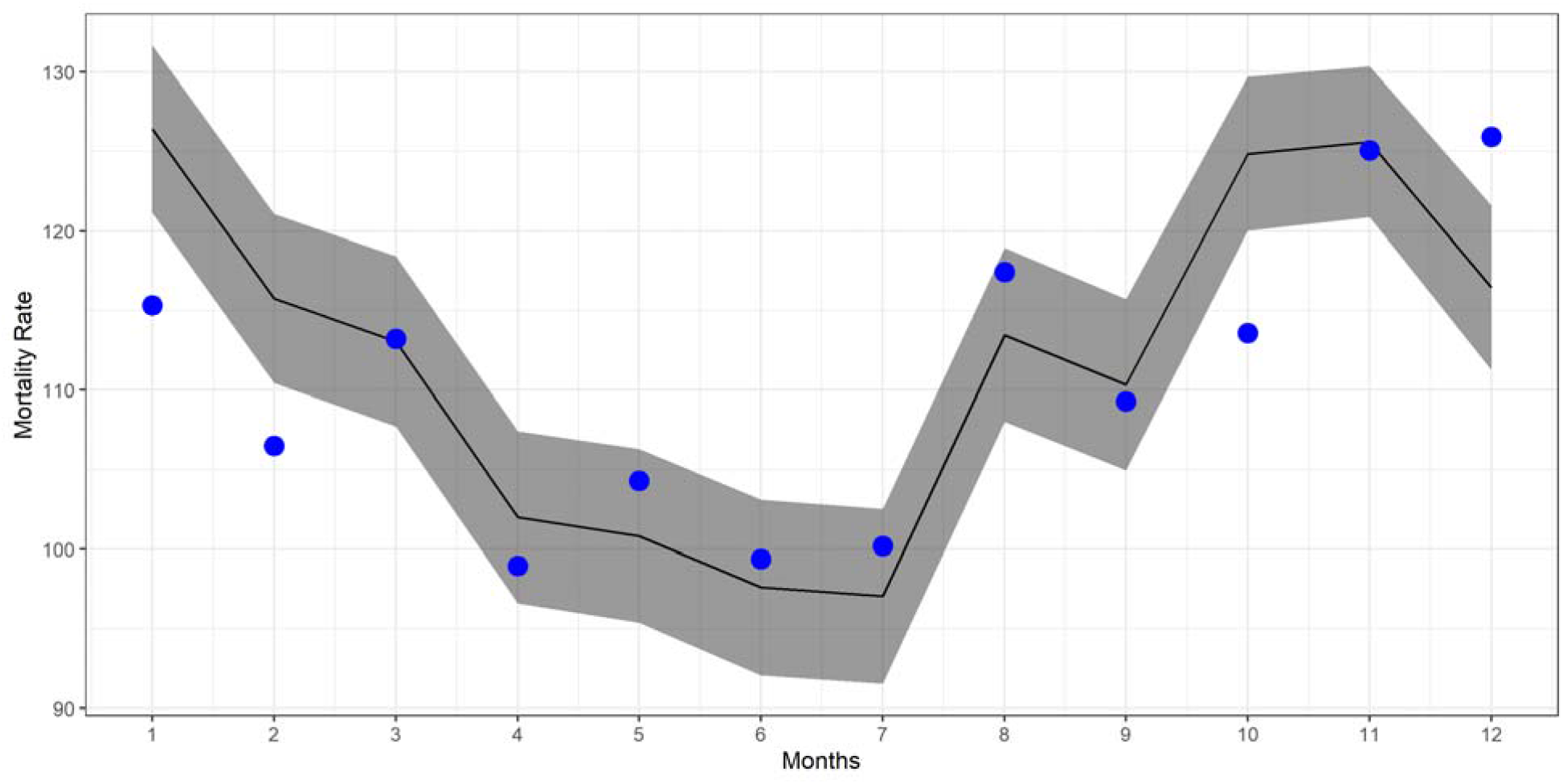
Predicted and observed all-cause mortality rates for females for the period March 2017 – February 2018 in Lithuania. Black lines indicate the forecast number of deaths without the 2017 taxation intervention, using the Lithuania population as standard. Blue dots indicate observed mortality rates.

